# NetSHy: Network Summarization via a Hybrid Approach Leveraging Topological Properties

**DOI:** 10.1101/2022.09.21.22280204

**Authors:** Thao Vu, Elizabeth M. Litkowski, Weixuan Liu, Katherine A. Pratte, Leslie Lange, Russell P. Bowler, Farnoush Banaei-Kashani, Katerina J. Kechris

## Abstract

Biological networks can provide a system level understanding of underlying processes. In many contexts, networks have a high degree of modularity, i.e., they consist of subsets of nodes, often known as subnetworks or modules, which are highly interconnected and may perform separate functions. In order to perform subsequent analyses to investigate the association between the identified module and a variable of interest, a module summarization, that best explains the module’s information and reduces dimensionality is often needed. Conventional approaches for obtaining network representation typically rely only on the profiles of the nodes within the network while disregarding the inherent network topological information. In this article, we propose NetSHy, a hybrid approach which is capable of reducing the dimension of a network while incorporating topological properties to aid the interpretation of the downstream analyses. In particular, NetSHy applies principal component analysis (PCA) on a combination of the node profiles and the well-known Laplacian matrix derived directly from the network similarity matrix to extract a summarization at a subject level. Simulation scenarios based on random and empirical networks at varying network sizes and sparsity levels show that NetSHy outperforms the conventional PCA approach applied directly on node profiles, in terms of recovering the true correlation with a phenotype of interest and maintaining a higher amount of explained variation in the data when networks are relatively sparse. The robustness of NetSHy is also demonstrated by more consistent correlation with the observed phenotype as the sample size decreases. Lastly, a genome wide association study (GWAS) is performed as an application of a downstream analysis, where NetSHy summarization scores on the biological networks identify more significant single nucleotide polymorphisms (SNP) than the conventional network representation.

## Introduction

Complex diseases are rarely a consequence of an abnormality on one single molecule, but rather the result of complex interactions and perturbations involving large sets of molecular components, which gives rise to the emergence of network-based approaches to gain a system-level understanding of the underlying biological processes (1). In particular, the informative patterns revealed by biological networks have been employed to gain insights into disease mechanisms (2), study comorbidities (3), facilitate therapeutic drugs and their targets (4), and discover network associated biomarkers (5). For instance, (6) generated a network consisting of 118 genes, in which a novel candidate gene, hyaluronan mediated motility receptor (HMMR), was demonstrated to associate with higher risk of breast cancer in humans. In another study, (7) constructed shared gene networks to uncover key drivers for cardiovascular disease (CVD) and type 2 diabetes (T2D), which in turn offered important insights for the development of therapeutic avenues targeting both diseases simultaneously.

Network analysis simplifies the complex biological systems to constituents (nodes) and their interactions (edges). Networks can be constructed directly based on gene expression data such as transcriptional regulatory networks (8) and coexpression networks (9), or can be built using the integration of multi-omics data (10). For example, in protein-protein interaction (PPI) networks, nodes are individual proteins and pairwise physical interactions are characterized by edges. Similarly, in co-expression networks, genes serve as nodes and their corresponding connecting edges are defined by the correlation between expression patterns. Utilizing the integration of multi-omics data, (11) captured the relationships between all pairs of transcripts and metabolites through a transcriptome-metabolome network.

While biological global networks provide a big picture of the underlying cellular processes, they are often too large to be considered as a whole. It has been shown that molecular networks have a high degree of modularity, i.e., they consist of subsets of nodes which are highly interconnected and may perform separate functions (12). Such collections of nodes are often known as modules or subnetworks. (13), for example, focused on studying the metabolic networks of 43 distinct organisms to uncover the hierarchical modularity property, which was shown to closely overlap with known metabolic functions. Additionally, acknowledging the advantages of network modular structure (14) regarding evolvability and robustness, (15) launched the community-driven challenge promoting assessment of different methods in identifying disease-relevant modules across a diversity of network types such as protein-protein interaction, homology and cancer-gene networks. Once identified, the subnetworks are related to external information in downstream analyses to obtain biologically meaningful interpretations. For instance, individual genes in a key module were modelled simultaneously in a LASSO-Cox regression framework to identify signature genes which were predictive of overall survival of patients with lung adenocarcinoma (LUAD) (16). Conversely, one can summarize a module into a feature which best explains the module’s behavior, referred to as a module representation, which then serves as direct input for downstream analyses. Such module centric approaches allow the collective impact of all entities in the identified module on an outcome of interest to be investigated (17, 18).

Denote a molecular profile of *n* subjects and *p* features as *X*_*n*×*p*_, a network highlighting the relationships between the features is represented by an adjacency matrix *A*_*p*×*p*_. Fig. S1 (Supplementary Information) outlines existing approaches for network representation, which utilize either *X*_*n*×*p*_ or *A*_*p*×*p*_. In the popular weighted correlation network analysis (WGCNA), (9) represented the gene expression profiles of a given network by the first principal component of *X*_*n*×*p*_, namely “eigengene”, denoted as *Z*_*n*×1_. The “eigengene” can be thought of as a weighted average expression of all individual genes in the network, in which the corresponding weights are defined such that the resulting “eigengene” can explain the most variation in the data. Similarly, (18) summarized each metabolite modules into “eigenmetabolite”, i.e., the first principal component of the network metabolic profile. The “eigenmetabolite” was subsequently used to identify significant genetic associations to make inferences about shared biochemical pathways. Alternatively, one can use the molecular profile of the most highly connected intramodular node, known as a hub node, as the network representation (19). The rationale for this approach is that hub nodes are more relevant to the functionality of networks than other nodes since they are central of the network’s architecture. For instance, in protein knockout experiments, hub proteins were shown to be essential for survival in lower organisms (17). While these approaches are capable of summarizing networks at the subject level, neither of them fully takes advantage of network topological properties. Specifically, the “eigengene” approach only focuses on the direction maximizing the variation in the measurements associated with nodes in the network which does not necessarily reflect the underlying connectivity structure between nodes. The “hub node” approach, on the other hand, projects the whole module information onto the profile of the single node with most connections while disregarding the roles of the remaining entities.

Networks are often defined as graphs from the graph theory perspective. There exist many graph embedding techniques which are designed to learn the graph topology directly using network adjacency matrix *A*_*p*×*p*_. In particular, matrix factorization based graph embeddings such as Graph Laplacian eigenmaps, multidimensional scaling (MDS) (20), and Isomap (21) exploited the network topology to create an interpoint distance matrix on which spectral decomposition was performed to extract a representation capturing the network structure at the node level. Furthermore, the emergence of deep learning in graph data has widened the scope of graph representation techniques. Deep learning based methods such as DeepWalk (22) and node2vec (23) deployed the truncated random walks (24), which were essentially the sets of paths sampled from the input graph to maximize the co-occurrence probability of the observing node’s neighborhood. In a different manner, autoencoders and deep neural networks can be applied directly to the proximity matrix of the whole graph rather than following random walk paths. More specifically, graph autoencoder (25) approaches such as structural deep network embedding (SDNE) (26) and sparse autoencoder (SAE) (27) minimized the reconstruction error of the representation output and the network input through encoder and decoder steps, such that nodes with similar neighborhood would have similar embeddings. The survey by (28) comprehensively reviewed each of these methods.

The aforementioned embedding techniques show promising results regarding reducing the dimension of input graphs while preserving topology information at the node level, i.e., transforming *A*_*p*×*p*_ to *Z*_*p*×*d*_ such that *d < p*. However, typical analyses linking module-specific features to clinical traits of interest (e.g., disease status, survival time, etc.) require a subject-level representation, i.e., *Z*_*n*×*d*_, with *d < p*. That becomes our motivation to propose an approach, NetSHy, that is capable of summarizing a network at a subject level while capturing the network topological properties. Specifically, NetSHy creates a latent matrix by combining the feature profile matrix *X*_*n*×*p*_ and network topology stored in a Laplacian matrix *L*_*p*×*p*_ prior to performing a principal component analysis (PCA) to obtain a summarization score for each subject. NetSHy is evaluated using inferred biological networks from a study on chronic obstructive pulmonary disease (COPD) (29) as well as simulated networks at different levels of network sparsity. The performance of the proposed approach, NetSHy, is compared to the conventional approach of just using the molecular profile without network information, which we refer to as NoNet, based on: (i) correlation with continuous phenotype and (ii) the variance of data explained by the resulting network summarization. We find that NetSHy outperforms NoNet in recovering the true correlation with the phenotype and maintaining a higher level of explained variation in the data when the networks are relatively sparse. Furthermore, NetSHy is proved to be more robust than NoNet when the sample size of the biological networks decreases. Lastly, we illustrate an example of a downstream analysis by performing a genome wide association study (GWAS) using the results of the network summarization and find that there are stronger signals when using network information through NetSHy compared to NoNet.

## Materials and Methods

### NetSHy

A weighted, non-negative, undirected network of *p* nodes can be represented by an adjacency matrix 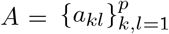, where *a*_*kl*_ reflects the similarity between nodes *k* and *l* in the network. Denote the corresponding feature profile of all nodes in the network as *X*_*n*×*p*_ with *n* and *p* representing numbers of subjects and features, respectively. Direct connection between any two nodes in the network can be reflected using the Laplacian matrix (30) as

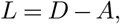

where, *A*_*p*×*p*_ is defined as above, *D* is a diagonal degree matrix such that *D*_*kk*_ *=*Σ*a*_*kl*_, *k, l =*1, …, *p*. The symmetric Laplacian matrix *L*_*p*×*p*_ records the direct connection of any two nodes as well as the node degree distribution in the network.

With *L*_*p*×*p*_ capturing the network topology, we define *X* ^*^, a transformation of *X*, as a combination of both node feature profiles and network topology, such that

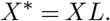

We then perform principal component analysis (PCA) on *X*^*^ to extract the first principal component (PC) of dimension *n*×1, as a representation capturing the variability in both directions of feature data and topology. For the rest of the paper, we will refer to the first PC obtained from *X* and *X*^*^ as NoNet and NetSHy summarization, denoted as *Z*_*NoNet*_ and *Z*_*NetSHy*_, respectively. Simultaneously, the corresponding first eigenvectors of size *p*×1, which store the direction and relative contribution of each node in the network to the summarizations, are denoted as *ϕ*_*NoNet*_ and *ϕ*_*NetSHy*_, respectively.

### Simulation scenarios

Two simulation scenarios were designed to evaluate the performance of NoNet and NetSHy summarization (columns (1) and (2) of Fig. S2).

- *Scenario (1):* Given a number of nodes *p* and a graph sparsity *α*_0_ ∈[0, 1], a network, denoted as *A*, was generated from a random model Renyi-Erdos (31) such that the probability of a node connecting to another node within a network was approximately *α*_0_. This was accomplished using the `R` package `igraph` (32). The edge weights 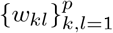 connecting nodes *k* and *l* were simulated from the uniform distribution such that *w*_*kl*_ ∼Unif(0.1, 0.8). Three network sizes of *p* = 30, 60, and 100, and three levels of sparsity *α*_0_ = 0.3, 0.6, and 0.9 were included in the study.
- *Scenario (2):* An adjacency matrix *A* was obtained directly from a previously published metabolite-protein network by Mastej et al. (29). Different from Scenario (1), the network size and sparsity level were fixed at *p* = 20 and *α*_0_ = 0.51, respectively. However, by applying hard-thresholding to remove weak edges, we were able to additionally assess the impact of network sparsity at *α*_0_ = 0.25.

Once the network adjacency matrix *A* was obtained, each offdiagonal element of the symmetric matrix *A* can be thought of a conditional relationship between the two corresponding features. According to a Gaussian graphical model, *A* served as a precision matrix Σ^−1^ to simulate the feature data, with additional estimation steps demonstrated in (33) to ensure Σ^−1^ a positive definite matrix. The feature data matrix *X*_0_ was generated such that *X*_0_ *∼ N* (0, Σ). The phenotype vector *Y*_0_ of size *n*×1 was obtained as a linear function of *X*_0_, as *Y*_0_ = *β*_0_ + *X*_0_*β* + *ϵ* with 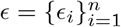 and 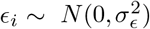. With the assumption that important nodes (i.e., nodes with high connectivity) had a large influence on the phenotype, we set *β*_0_ *∼ N* (0, 1), and 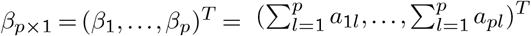, with *a*_*kl*_ as the (*kl*)th element of the adjacency matrix *A*. We further extended the simulation by perturbing the true data matrix to obtain the observed data matrix *X* such that *X* = *X*_0_ + *E* where 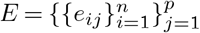 and 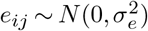 denoted the noise matrix. This simulation setup is to mimic the real contexts, as follows. Given a subnetwork of size *p*, we obtain the corresponding adjacency matrix *A*_*p*×*p*_ by directly subsetting the global network instead of re-estimating it (say, 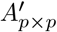 using the feature profile *X*_*n*×*p*_ of only the nodes within the subnetwork. In other words, *A*_*p*×*p*_ and 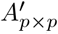 are different in a sense that *A* captures the global signals shared by all features in the dataset while *A*^*′*^only reflects local signals. Sequentially, *X*_0_ (true profile) of dimension *n*×*p* and *X*_*n*×*p*_ (observed profile) are induced from *A* and *A*^*′*^, respectively. However, in reality, we only observe *X*_*n*×*p*_ which can be thought of as the feature profile being contaminated with measurement errors. By leveraging the network information inherent in *A*_*p*×*p*_, we would expect to recover some true underlying signals which might have been lost due to such measurement perturbations.

Furthermore, across the two scenarios, we rigorously investigated the impact of sample size on each method performance. More specifically, we started at the sample size of *n* = 1000 subjects, and random subsamplings were iterated 1000 times for each sample size of 500, 300, 200, 100, and 50, respectively, to evaluate the robustness of each summarization regarding both criteria detailed in the next section. Note that at each sample size except for *n* = 1000, mean and standard deviation were calculated from the 1000 iterations.

#### Evaluation criteria

With the available observed data matrix *X*_*n*×*p*_, phenotype vector *Y*_0_ of size *n*×1, and the network adjacency matrix *A*_*p*×*p*_, we obtained the subject-level score vectors using NoNet and NetSHy approaches (Section), denoted as *Z*_*NoNet*_ and *Z*_*NetSHy*_, respectively. We then evaluated the performance of the two scores using the following criteria:

- Correlation of each summarization score with the true phenotype *Y*_0_ was calculated as:

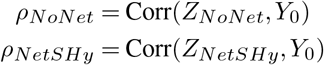
- Proportion of variance of the true feature profiles *X*_0_ explained (PVE) by each of the two summarization versions NoNet and NetSHy, using the associated first eigenvectors *ϕ*_*NoNet*_ and *ϕ*_*NetSHy*_, respectively was computed as follows:

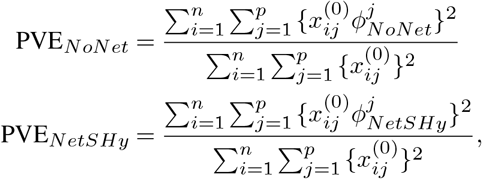

with 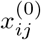 as the (*ij*)th element of 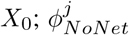 as the *j*th element of *ϕ*_*NoNet*_; and 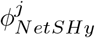 as the *j*th element of *ϕ*_*NetSHy*_.

The two quantities above were compared to the optimal correlation and PVE, denoted as *ρ*_*opt*_ and PVE_*opt*_, respectively, which were computed directly from the true data matrix *X*_0_. In particular, the first PC and first eigenvector, denoted as *Z*_*opt*_ and *ϕ*_*opt*_ respectively, obtained from *X*_0_ were used to compute *ρ*_*opt*_ and PVE_*opt*_, as follows:

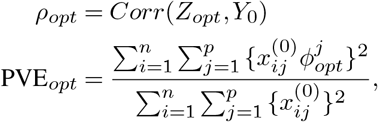

with 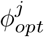 as the *j*th element of *ϕ*_*opt*_. The closer the values to *ρ*_*opt*_ and PVE_*opt*_, the better the performance.

### Biological networks

The applicability of NetSHy was further validated using biological networks (column (3) of Fig. S2) specific for chronic obstructive pulmonary disease (COPD), regarding performance robustness and interpretable results. We used a metabolite-protein (M-P) network for robustness assessment and a protein (P) network as a GWAS use case of the method. Note that the observed data *X* and phenotype *Y* were used for the analysis directly without any simulation involved.

#### COPDGene and COPD Phenotype

The COPDGene study is a multi-center study that enrolled 10,198 participants including non-Hispanic whites and African Americans with and without COPD between 2007 and 2011 (Visit 1). Five-year follow up visits took place from 2013 to 2017 (Visit 2). Study participants from Visit 2, after removing individuals with lung transplant or lung reduction surgery and never smokers, provided consent; and their blood samples were used for-omic analyses.

COPD was defined by spirometric evidence of airflow obstruction, which was computed as a ratio of postbronchodilator forced expiratory volume at one second (FEV1) to forced vital capacity (FVC). FEV1% is the amount of air one can forcibly exhale in one second divided by the predicted FEV1 adjusted for age, height, race, and sex (34). The Global Obstructive Lung Disease (GOLD) system is used to grade COPD. More information on the GOLD system can be found in Section S.2 (Supplementary Information).

#### COPDGene Genotyping

COPDGene subjects were of selfreported non-Hispanic white (NHW) or African-American ancestry, and genotyped as previously described by Cho et al. (35). Briefly, genotyping was performed using the HumanOmniExpress array, and BeadStudio quality control, including reclustering on project samples was performed following Illumina guidelines. Subjects and markers with a call rate of *<* 95% were excluded. Population stratification exclusion and adjustment on self-reported white subjects was performed using EIGENSTRAT (EIGENSOFT Version 2.0).

#### Proteomic data

The following two platforms were used to quantify proteomic data in Visit 2 of COPDGene. <u>*SOMAScan v1*.*3:*</u> P100 plasma was profiled using SOMAscan^®^ Human Plasma 1.3K assay (SomaLogic, Boulder, CO, USA) at National Jewish Health. SOMAScan is a multiplex proteomic assay quantified by microarrays. This assay measured 1317 SOMAmers which are short single-stranded deoxyoligonucleotides (aptamers) binding with high affinity and specificity to specific protein structures (36). <u>*SOMAScan v4*.*0:*</u> This SOMAScan platform used 4,979 different SOMAmers to quantify 4,776 unique proteins with 4,720 unique Uniprot numbers. Details on the preprocessing steps of the proteomic data are given in Section S.2 (Supplementary Information).

#### Metabolomic data

The same P100 plasma was profiled using the Metabolon (Durham, NC, USA) Global Metabolomics platform to quantify 1392 metabolites. After filtering for missing values, 995 metabolites were used in the analysis. More details can be found in Section S.2 (Supplementary Information).

#### Metabolite - protein (M-P) network construction

We used a subset of the COPDGene participants who had both metabolomic and proteomic data available at Visit 2 to construct a M-P network via Sparse Multiple Canonical Correlation Network (SmCCNet) introduced by (37). The two - omic data were adjusted for white blood cell count, percent eosinophil, percent lymphocytes, percent neutrophiles, and hemoglobin as these covariates may influence metabolite and protein abundance in human blood studies. Then, SmCCNet was applied to the adjusted metabolomic (*p*_1_ = 995 metabolites) and proteomic (*p*_2_ = 1317 proteins) data to construct multi-omic networks correlated with the phenotype FEV1% (*n* = 994 subjects) via multiple canonical correlation approach. In essence, SmCCNet maximized the correlation between the two omics datasets (i.e., metabolomics and proteomics) and the phenotype FEV1% while imposing a sparsity to de-emphasize the impacts of metabolites and proteins which did not contribute to the overall correlation. After hierarchical clustering and hard thresholding to filter out weak edges, strongly connected subnetworks that were well correlated with FEV1% were identified. More details can be found in (29). In this work, we used a M-P network for FEV1% consisting of seven metabolites and thirteen proteins.

### Robustness assessment

We assume that the collected metabolomics and proteomics data (i.e., *X*) are perturbed measurements due to instrument error of the true but non-observed metabolite and protein (i.e., *X*_0_) levels. With *X*_0_ not available, the comparison of NetSHy and NoNet relative to the optimal level (Section) was not obtainable. We instead focused on assessing the robustness of the two approaches regarding the correlation with the observed phenotype as the sample size decreased. The observed data corresponding to the identified M-P network *X*_994×20_ and phenotype *Y*_994×1_ were randomly sampled at decreasing sizes: 500, 300, 200, 100, and 50, respectively, and repeated for 1000 iterations. Mean and standard deviation of the correlation of each summarization: *Z*_*NetSHy*_, *Z*_*NoNet*_, with the observed phenotype were recorded at each sample size, except for the full sample size *n* = 994. The robust performances of NetSHy and NoNet were assessed using the dropping rates as the sample size decreased.

#### Protein (P) network construction

We used a subset of the COPDGene non-Hispanic white participants with proteomic data available at Visit 2 to construct protein networks. The proteomic data collected from SOMAScan v4.0 platform (Section) had larger sample size to perform a GWAS since the data did not need to be matched with the metabolomic data. Similar to M-P network construction, SmCCNet ((37) was applied to the proteomic data (*p* = 4776 proteins) to construct protein (P) networks maximizing correlation with the phenotype FEV1% (*n* = 1660 subjects). A 5-fold cross validation was used on a set of sparsity parameters from 0.1 to 0.5 with a step size of 0.1 to select an optimal value that minimized the prediction error. After hierarchical clustering and weak edge trimming, a strongly connected network of sixteen proteins and well correlated with FEV1% was identified.

### GWAS analysis

Genome-wide association study (GWAS) analysis was used to demonstrate the applicability of the summarization methods (i.e., NetSHy and NoNet) in an example downstream analysis. Specifically, by applying each approach to the identified protein network (*X*_1660×16_, *Y*_1660×1_), we obtained summarization scores, *Z*_*NetSHy*_ and *Z*_*NoNet*_. The resulting summarization scores were inverse-normalized prior to linearly regressing on the genotype while adjusting for covariates including age, BMI, gender, smoking status, and five genetic principal components (PCs) (38). The genetic PCs were obtained from previously performed analysis including only COPDgene participants (35). In total, there were 14,553,332 variants tested for significant association with the protein network across the subjects. Section S.2 (Supplementary Information) includes our detailed GWAS analysis.

## Results

### Simulation results

Fig. 1 depicts the performances of NetSHy and NoNet summarization scores in terms of correlation (*ρ*) with true phenotype (top row) and proportion of variance explained (PVE) in true data matrix *X*_0_ (bottom row) with respect to the optimal quantities *ρ*_*opt*_ and PVE_*opt*_, respectively. The closer the values to 1, the better the performances. In addition, the network size was fixed at *p* = 30 while the level of sparsity increased from *α*_0_ = 0.3 to *α*_0_ = 0.9. Across the three sparsity levels, both approaches deviated from the optimal level as the sample size decreased. In general, the trend was observed that NoNet dropped at a faster rate compared to NetSHy. Interestingly, NetSHy had higher PVE in comparison with NoNet regardless of sample size and network sparsity. However, as the nodes were connected more densely, i.e., larger *α*_0_, the deviation in PVE between the two approaches became less apparent. More specifically, at *α*_0_ = 0.3 and *n* = 50, the ratio of PVE_*NetSHy*_ to the optimal PVE_*opt*_ was 0.81 while such ratio of NoNet was 0.57. However, when *α*_0_ = 0.9, the PVEs of NetSHy and NoNet with respect to the optimal PVE were 0.66 and 0.56, respectively. A similar pattern was observed for *ρ*, but at a more subtle level. For instance, at *α*_0_ = 0.3 and *n* = 50, the correlations *ρ*_*NetSHy*_ and *ρ*_*NoNet*_ with respect to the optimal correlation *ρ*_*opt*_ were 0.83 and 0.75, respectively. When the sparsity level increased to *α*_0_ = 0.9, the ratio of *ρ*_*NetSHy*_ to *ρ*_*opt*_ was 0.72 while such ratio of NoNet was 0.67. Fig. S3 and Fig. S4 (Supplementary Information) show the same set of results for network size *p* = 60 and *p* = 100, respectively. Slightly different from the previous case, NetSHy still showed some improvement over NoNet in both *ρ* and PVE at sparsity level *α*_0_ = 0.3 when *p* = 100. However, in the case of a densely connected network at *α*_0_ = 0.9, NetSHy and NoNet performed almost identical.

**Fig. 1.**
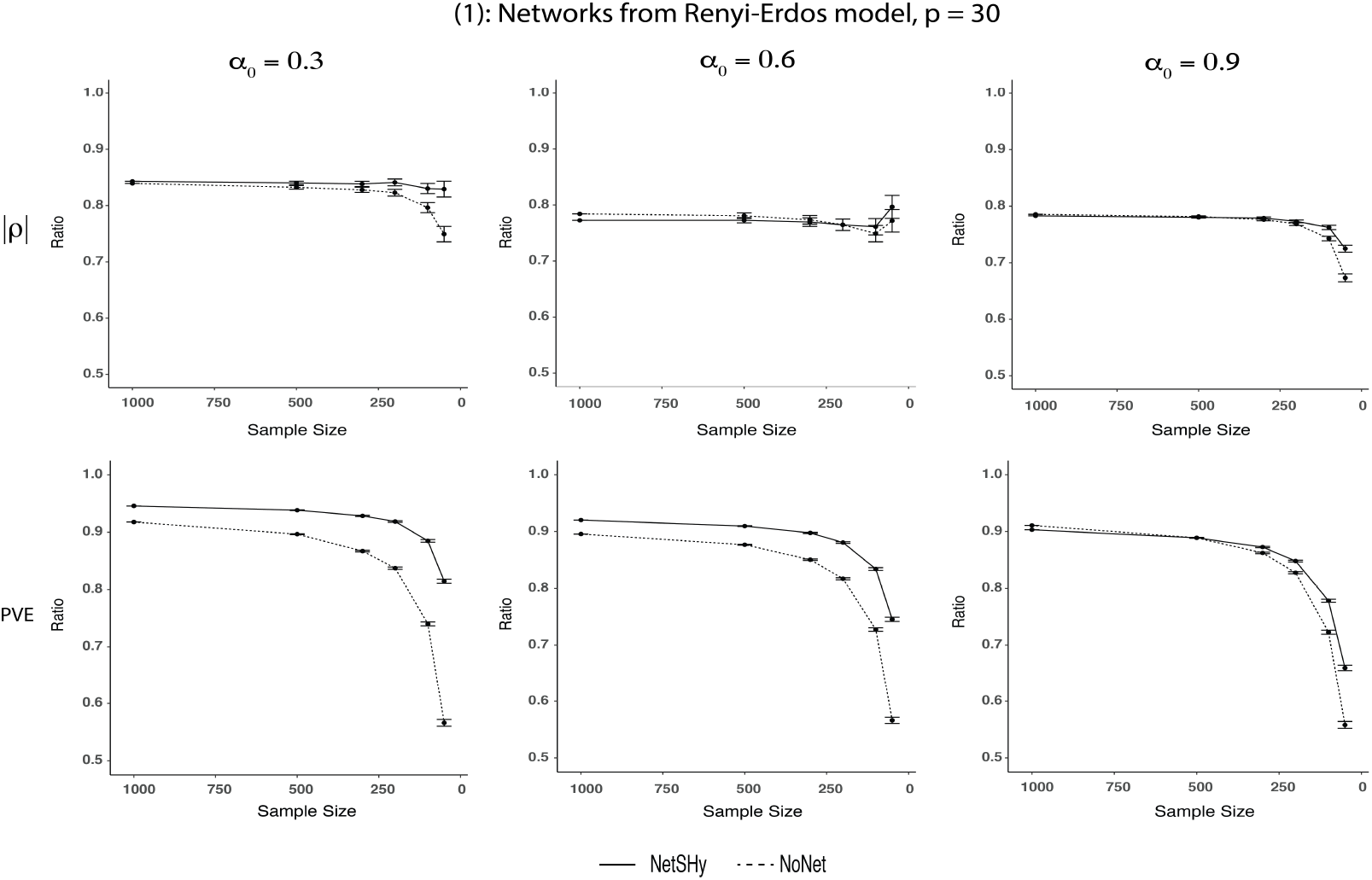
Results of Simulation Scenario (1), π= 30: Fixing network size at *p* = 30 while varying sparsity levels from *α*_0_ = 0.3 to *α*_0_ = 0.9, NetSHy and NoNet were assessed using correlation with phenotype (*ρ*) and proportion of variance explained (PVE) relative to the optimal level, as sample size decreased. Specifically, the sample size was started at 1000 subjects, and random subsamplings were iterated 1000 times for each sample size of 500, 300, 200, 100, and 50, respectively. The closer the value to 1, the better the performance. The error bars summarize the standard deviations of *ρ* and PVE from the 1000 iterations at each sample size except for *n* = 1000. The range of *ρ* and PVE ratio in the y-axis is between 0 and 1. However, we have zoomed in between 0.5 and 1 for better visualization.

Fig. 2 illustrates the performances of NetSHy and NoNet summarization scores in the empirical-based simulation (Simulation scenario (2) in Fig. S2) at two levels of network sparsity *α*_0_ = 0.25 and 0.51 across decreasing sample sizes. Similar to simulation scenario (1), the two approaches deviated from the optimal level as sample size decreased. Though at *α*_0_ = 0.25, NetSHy suffered a little in recovering true correlation with the phenotype at sample sizes of 1000 and 500, it was more robust at more extreme sizes of 100 and 50. Specifically, at *n* = 1000, the ratio of NetSHy correlation *ρ*_*NetSHy*_ to the optimal level *ρ*_*opt*_ was 0.63 while that ratio of NoNet was 0.68. However, at the smallest sample size of *n* = 50, the correlation of NoNet scores with the phenotype decreased greatly, which caused its ratio with the optimal correlation *ρ*_*opt*_ to drop to 0.47 while such ratio of NetSHy remained around 0.62. At *α*_0_ = 0.51, NetSHy and NoNet performed almost identical when sample sizes were large. In particular, at *n* = 1000, the ratios of NetSHy and NoNet correlations with respect to the optimal level were 0.66 and 0.67, respectively. The improvement of NetSHy over NoNet was more appreciable towards the extreme sizes of 100 and 50. More precisely, at *n* = 50, NetSHy maintained the correlation ratio to the optimal level at around 0.66 while that ratio using NoNet scores reduced to 0.55. Regarding PVE, NetSHy outperformed NoNet approach at all sample sizes. Similar to what had been observed, the improvement of NetSHy over NoNet was more substantial towards smaller sample sizes.

**Fig. 2.**
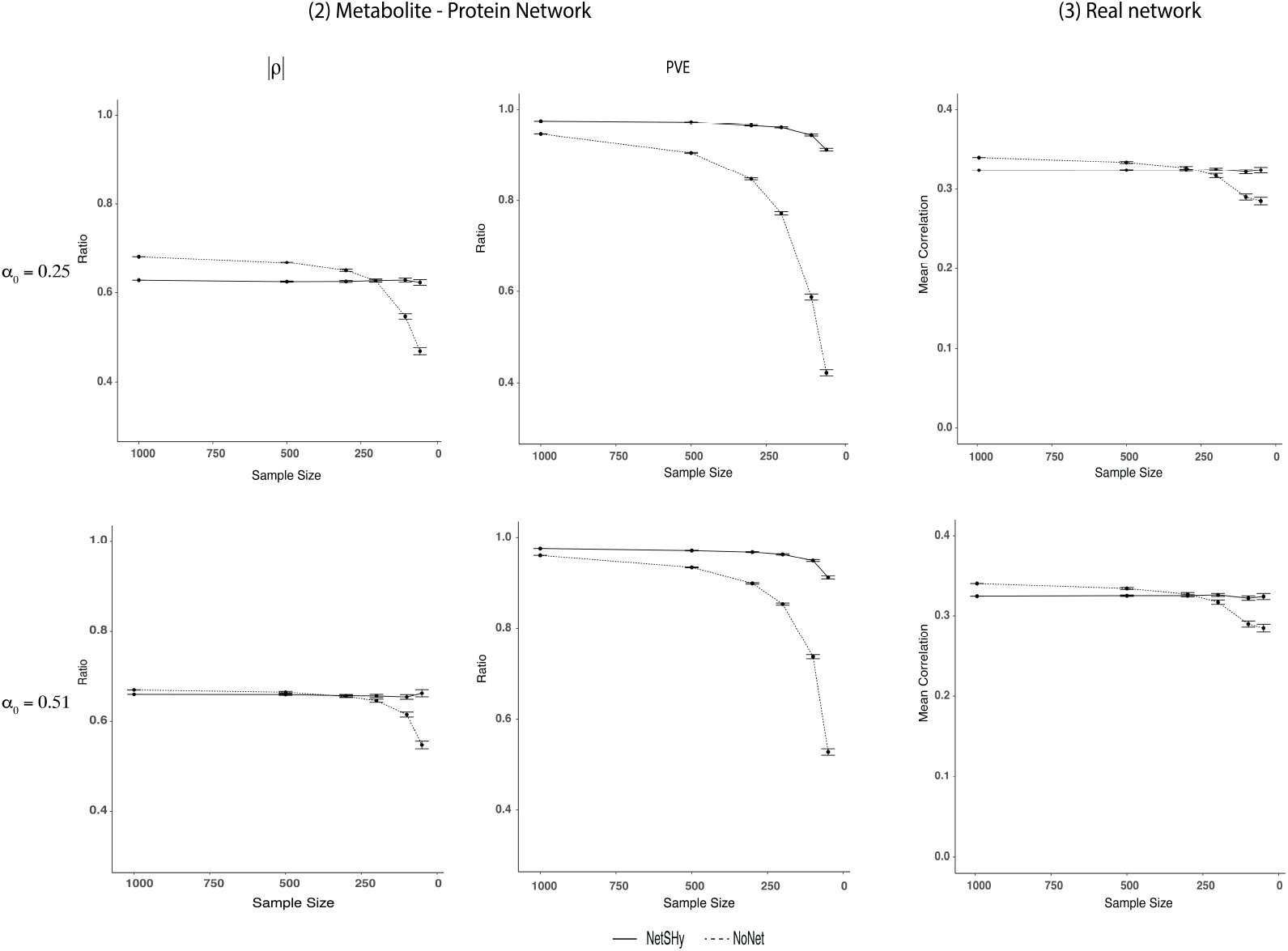
Results of Simulation Scenario (2) and Real Network (3): <u>*First two columns:*</u> Simulation based on a published metabolite-protein network with fixed *p* = 20 at two sparsity levels of *α*_0_ = 0.25 (top row) *α*_0_ = 0.51 (bottom row). NetSHy and NoNet were assessed using correlation with phenotype (*ρ*) and proportion of variance explained (PVE) relative to the optimal level, as sample size decreased. Specifically, the sample size was started at 1000 subjects, and random subsamplings were iterated 1000 times for each sample size of 500, 300, 200, 100, and 50, respectively. The closer the value to 1, the better the performance. The range of *ρ* and PVE ratio in the y-axis is between 0 and 1. However, we have zoomed in between 0.4 and 1 for better visualization. <u>*Last column:*</u> Published metabolite-protein network with fixed *p* = 20 at two sparsity levels of *α*_0_ = 0.25 (top) *α*_0_ = 0.51 (bottom) was used directly for evaluation. Without knowledge of true underlying data matrix *X*_0_, proportion of variance explained (PVE) was not assessed. Instead, the robustness of each approach regarding observed correlation with phenotype was of interest as the sample size decreased. Specifically, random subsamplings were iterated 1000 times for each sample size of 500, 300, 200, 100, and 50, respectively. The lower the dropping rate of a method’s trajectory, the more robust the performance. The error bars summarize the standard deviations of *ρ* from the 1000 iterations at each sample size except for *n* = 994.

### Application results

The evaluation of NetSHy and NoNet was further validated using biological networks, M-P and P networks, with respect to performance robustness and GWAS results.

### Robustness

Last column of Fig. 2 presents the mean correlation with observed phenotype *ρ* using the M-P network of size *p* = 20 at two sparsity levels *α*_0_ = 0.25 and *α*_0_ = 0.51. Similar patterns were observed across the two *α*_0_ levels that the correlation *ρ* dropped as the sample size got smaller. At the original sample size *n* = 994, the observed NoNet correlation with the phenotype (|*ρ*_*NoNet*_| = 0.34) was slightly higher than the corresponding NetSHy correlation (|*ρ*_*NetSHy*_| = 0.32). The difference between these two correlations was not significant with p-value of 0.49 using bootstrapping. Interestingly, the overall trajectory of NetSHy remained relatively stable even at the small sample size (*n* ≤100) while NoNet suffered a substantial drop. For instance, at *n* = 50 and *α*_0_ = 0.51, the mean correlation of NetSHy was 0.32 while that of NoNet reduced to 0.28.

During the subsampling process, as we selected 500 subjects out of 994 without replacement, the subjects across iterations overlapped to different degrees due to random chance. Intuitively, the overlap was greater for larger sample sizes, leading to less variation across iterations. As such, we observed larger computed standard deviations of correlation with phenotype (*ρ*) and proportion of variance explained (PVE) as we decreased sample size from *n* = 500 to *n* = 50 (Fig. 2). Similar patterns were also observed in the simulation studies (Fig. 1).

### GWAS results and interpretation

To demonstrate a downstream application of network summarization, we tested whether any single nucleotide polymorphisms (SNPs) had a significant association with the protein network across the subjects. This analysis would be useful for identifying potential regulators of the network. Fig. 3 (a), (b) show the GWAS results for NetSHy and NoNet, respectively. At a threshold level of 5×10−8, NetSHy identified twenty-four significant SNPs while NoNet detected only one. The top SNP, rs1017301 (Chromosome 12: 9210335), was discovered using NetSHy score (p = 2.38×10−13, MAF = 0.33) whereas the same SNP did not reach significance (p =4.26×10−4) using the NoNet approach. For NoNet summary score, the top SNP was rs118028480 (Chromosome 22: 39592172, p = 4.35×10−8). Section S.5 (Supplementary Information) provides a full interpretation of the significant SNPs obtained using NetSHy and NoNet summarization scores.

**Fig. 3.**
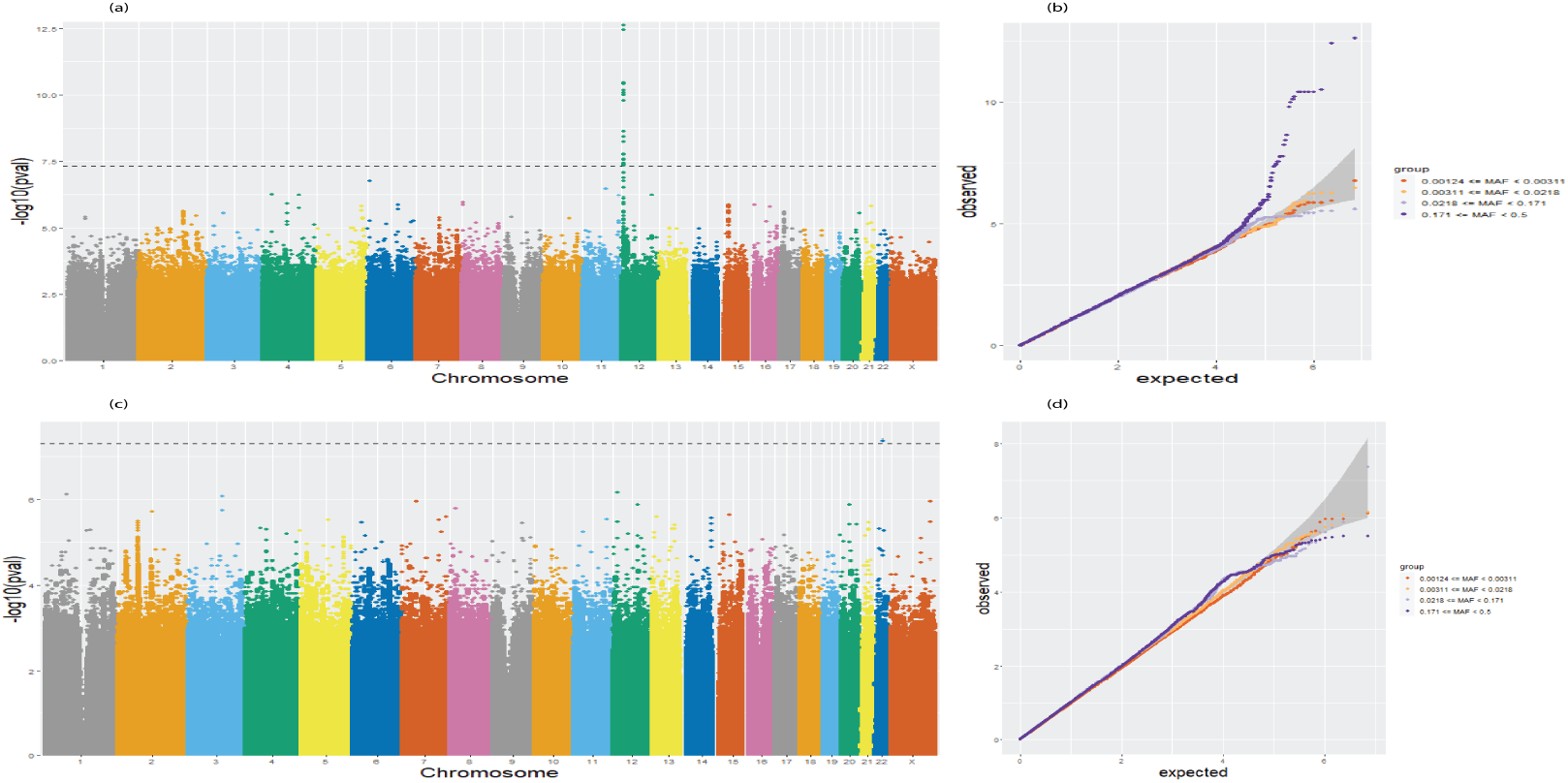
Results from GWAS studies on protein network specific to FEV1. **Top Row:** NetSHy summarization was used to linearly regress on genotype to identify significantly associated SNPs. (a) Manhattan plot using NetSHy summarization. (b) QQ plot using NetSHy summarization. **Bottom Row:** NoNet summarization was used to linearly regress on genotype to identify significantly asscoiated SNPs. (c) Manhattan plot using NoNet summarization. (d) QQ plot using NoNet summarization.

Recall that NetSHy summary score is a weighted average abundance of all proteins in the network with the relative weights determined by performing PCA on the combination of network topology and the corresponding node feature profiles (i.e., *X*^*\**^ in Section 2.1). Fig. S5 (Supplementary Information) shows weights of five proteins which contribute the most to the NetSHy summary score. The five proteins included Fructose-bisphosphate aldolase B (ALDOB), Fructose-1,6-bisphosphatase 1 (FBP1), Argininosuccinate lyase (ASL), Ferritin, and Ferritin light chain (FTL). The correlation between the NetSHy summary score and FEV1% is 0.36. Interestingly, by checking the correlation of each individual protein with FEV1%, we noticed that the absolute values of the correlation ranged from 0.14 to 0.30. In other words, by using a summary score as an aggregate of all proteins in the network, we saw an increase in the correlation with the phenotype. A detailed description of the relationship between the top proteins with lung diseases, particularly COPD, is given in Section S.5 (Supplementary Information).

## Discussion and Conclusions

Biological networks provide a system level understanding of the underlying cellular processes, but they are often too large to be considered as a whole. As a result, subsets of nodes (i.e., modules) which are highly connected to each other may be considered. Furthermore, a purpose of many network analyses is to relate the resulting modules to external sample information in downstream analyses, depending on the research question of interest. However, due to the multidimensional nature of networks, they need to be summarized prior to subsequent analyses. Conventional approaches rely on the feature profiles of the within-network entities while disregarding the inherent connectivity properties to obtain a network representation. As such, the summarization results do not truly reflect the roles of individual biological entities in the network. This motivates us to propose NetSHy, a hybrid approach which is capable of reducing the dimension of networks while accounting for both node profiles and topological properties. In our preliminary analysis (Section S.6, Supplementary Information), we explored two methods to incorporate topology in network summarization, i.e., a diffusion process (39, 40) and a weighted approach accounting for a secondary proximity embedded in a topology overlap matrix (TOM) (9). However, we did not pursue further with the comparisons due to instability or suboptimal results. Thus, we only compare the performance of NetSHy with NoNet (i.e., not including network information) through simulation scenarios based on random and empirical networks at varying levels of network size and sparsity, with regard to the ability to recover true correlation with the phenotype of interest and the amount of true variation explained. Furthermore, the robustness of the two approaches is assessed using biological networks via repeated subsamplings at a decreasing level. Finally, we validate the applicability of NetSHy and NoNet approach using the GWAS analysis.

NetSHy outperforms the NoNet approach regarding both correlation with true phenotype (*ρ*) and proportion of variance explained (PVE), when the networks are relatively small and sparse. However, when networks increase in size and the nodes are more densely connected, the improvement of NetSHy over NoNet is not as pronounced. This is not unexpected as when almost every node in the network is interconnected, the connectivity roles of individual nodes are similar. Thus, leveraging topological properties in this scenario might provide no additional gain for NetSHy, as compared to the NoNet approach. In applications on biological networks, the robustness of both approaches is brought into focus, due to lacking true underlying relationship between phenotype and feature data. In the M-P network, the observed correlation with the COPD phenotype FEV1% of NetSHy is slightly lower than that of NoNet at full sample size (0.34 vs. 0.32). Though the difference was insignificant (p = 0.49), it is still worth noting. However, by random subsampling at reducing sizes, NetSHy’s trajectory of observed correlation with the phenotype drops at a slower rate compared to NoNet, indicating NetSHy is more robust to small sample sizes. Finally, in the GWAS analysis of a protein network, NetSHy and NoNet summarization scores are used as response variables in a linear regression framework with genotype and other covariates. NetSHy identifies more significant SNPs associated with a given network, compared to NoNet approach.

We have presented promising results of NetSHy in representing networks at the subject level, however, we have still relied on the linearity assumption of the classical dimension reduction PCA. Additionally, the topological properties stored in the Laplacian matrix might not be sufficient for capturing the local neighborhood structure when the networks grow bigger and/or denser, as seen in the “large *p*, large *α*_0_” simulation scenario. We could potentially leverage the Isomap approach (21) to modify *X*^*\**^. Particularly, for any two nodes in a network, instead of their direct connection, the geodesic distances computing their shortest path distances could be used to represent the connectivity measure. Such connectivity matrix would then replace the *L* matrix in the calculation of *X*^*\**^. Lastly, kernel PCA (41) could be applied on *X*^*\**^ to extract the low-dimensional nonlinear representation. Alternatively, we have considered different techniques extracting information contained in large protein - protein interaction (PPI) networks such as FUSE (42), VoG (43), GraSS (44), SNAP and k-SNAP (45), CANAL (46). However, the summaries acquired from these approaches are themselves graphs. We could potentially use these approaches in place of the thresholding counterpart to simultaneously trim off weak edges and simplify the networks prior to summarizing them. This work is currently under investigation.

## Data Availability

All data produced in the present study are available upon reasonable request to the authors.

## Acknowledgements

The COPDGene^®^ project is supported by the COPD Foundation through contributions made to an Industry Advisory Board comprised of AstraZeneca, Boehringer-Ingelheim, Genentech, GlaxoSmithKline, Novartis, and Sunovion.

## Bibliography

1. Giorgio Valentini, Alberto Paccanaro, Horacio Caniza, Alfonso E Romero, and Matteo Re. An extensive analysis of disease-gene associations using network integration and fast kernel-based gene prioritization methods. Artificial Intelligence in Medicine, 61(2):63–78, 2014.

2. Peng Zhang and Yuval Itan. Biological network approaches and applications in rare disease studies. Genes, 10(10):797, 2019.

3. Jessica Xin Hu, Cecilia Engel Thomas, and Søren Brunak. Network biology concepts in complex disease comorbidities. Nature Reviews Genetics, 17(10): 615–629, 2016.

4. Giulia Fiscon, Federica Conte, Lorenzo Farina, and Paola Paci. Network-based approaches to explore complex biological systems towards network medicine. Genes, 9(9):437, 2018.

5. Tuba Sevimoglu and Kazim Yalcin Arga. The role of protein interaction networks in systems biomedicine. Computational and structural biotechnology journal, 11(18):22–27, 2014.

6. Miguel Angel Pujana, Jing-Dong J Han, Lea M Starita, Kristen N Stevens, Muneesh Tewari, Jin Sook Ahn, Gad Rennert, íctor Moreno, Tomas Kirchhoff, Bert Gold, et al. Network modeling links breast cancer susceptibility and centrosome dysfunction. Nature genetics, 39(11):1338–1349, 2007.

7. Le Shu, Kei Hang K Chan, Guanglin Zhang, Tianxiao Huan, Zeyneb Kurt, Yuqi Zhao, Veronica Codoni, David-Alexandre Trégouët, Cardiogenics Consortium, Jun Yang, et al. Shared genetic regulatory networks for cardiovascular disease and type 2 diabetes in multiple populations of diverse ethnicities in the united states. PLoS genetics, 13(9):e1007040, 2017.

8. Xiaohui Chen, Ming Chen, and Kaida Ning. Bnarray: an r package for constructing gene regulatory networks from microarray data by using bayesian network. Bioinformatics, 22(23):2952–2954, 2006.

9. Bin Zhang and Steve Horvath. A general framework for weighted gene coexpression network analysis. Statistical applications in genetics and molecular biology, 4(1), 2005.

10. Johann S Hawe, Fabian J Theis, and Matthias Heinig. Inferring interaction networks from multi-omics data. Frontiers in genetics, 10:535, 2019.

11. Jörg Bartel, Jan Krumsiek, Katharina Schramm, Jerzy Adamski, Christian Gieger, Christian Herder, Maren Carstensen, Annette Peters, Wolfgang Rathmann, Michael Roden, et al. The human blood metabolome-transcriptome interface. PLoS genetics, 11(6):e1005274, 2015.

12. Roger P Alexander, Philip M Kim, Thierry Emonet, and Mark B Gerstein. Understanding modularity in molecular networks requires dynamics. Science signaling, 2(81):pe44–pe44, 2009.

13. Erzsébet Ravasz, Anna Lisa Somera, Dale A Mongru, Zoltán N Oltvai, and A-L Barabási. Hierarchical organization of modularity in metabolic networks. science, 297(5586):1551–1555, 2002.

14. Gustavo Caetano-Anollés, M Fayez Aziz, Fizza Mughal, Frauke Gräter, Ibrahim Koç, Kelsey Caetano-Anollés, and Derek Caetano-Anollés. Emergence of hierarchical modularity in evolving networks uncovered by phylogenomic analysis. Evolutionary Bioinformatics, 15:1176934319872980, 2019.

15. Sarvenaz Choobdar, Mehmet E Ahsen, Jake Crawford, Mattia Tomasoni, Tao Fang, David Lamparter, Junyuan Lin, Benjamin Hescott, Xiaozhe Hu, Johnathan Mercer, et al. Assessment of network module identification across complex diseases. Nature methods, 16(9):843–852, 2019.

16. Yuan Wu, Lingge Yang, Long Zhang, Xinjie Zheng, Huan Xu, Kai Wang, and Xianwu Weng. Identification of a four-gene signature associated with the prognosis prediction of lung adenocarcinoma based on integrated bioinformatics analysis. Genes, 13(2):238, 2022.

17. Peter Langfelder, Paul S Mischel, and Steve Horvath. When is hub gene selection better than standard meta-analysis? PloS one, 8(4):e61505, 2013.

18. Pascal Schlosser, Yong Li, Peggy Sekula, Johannes Raffler, Franziska Grundner-Culemann, Maik Pietzner, Yurong Cheng, Matthias Wuttke, Inga Steinbrenner, Ulla T Schultheiss, et al. Genetic studies of urinary metabolites illuminate mechanisms of detoxification and excretion in humans. Nature genetics, 52(2):167–176, 2020.

19. Peter Langfelder and Steve Horvath. Wgcna: an r package for weighted correlation network analysis. BMC bioinformatics, 9(1):1–13, 2008.

20. Thomas Hofmann and Joachim Buhmann. Multidimensional scaling and data clustering. Advances in neural information processing systems, pages 459–466, 1995.

21. Joshua B Tenenbaum, Vin de Silva, and John C Langford. A global geometric framework for nonlinear dimensionality reduction. science, 290(5500):2319–2323, 2000.

22. Bryan Perozzi, Rami Al-Rfou, and Steven Skiena. Deepwalk: Online learning of social representations. In i>Proceedings of the 20th ACM SIGKDD international conference on Knowledge discovery and data mining, pages 701–710, 2014.

23. Aditya Grover and Jure Leskovec. node2vec: Scalable feature learning for networks. In Proceedings of the 22nd ACM SIGKDD international conference on Knowledge discovery and data mining, pages 855–864, 2016.

24. Frank Spitzer. Principles of random walk, volume 34. Springer Science & Business Media, 2013.

25. Pascal Vincent, Hugo Larochelle, Isabelle Lajoie, Yoshua Bengio, Pierre-Antoine Manzagol, and Léon Bottou. Stacked denoising autoencoders: Learning useful representations in a deep network with a local denoising criterion. Journal of machine learning research, 11(12), 2010.

26. Daixin Wang, Peng Cui, and Wenwu Zhu. Structural deep network embedding. In Proceedings of the 22nd ACM SIGKDD international conference on Knowledge discovery and data mining, pages 1225–1234, 2016.

27. Fei Tian, Bin Gao, Qing Cui, Enhong Chen, and Tie-Yan Liu. Learning deep representations for graph clustering. In Proceedings of the AAAI Conference on Artificial Intelligence, volume 28, 2014.

28. Hongyun Cai, Vincent W Zheng, and Kevin Chen-Chuan Chang. A comprehensive survey of graph embedding: Problems, techniques, and applications. IEEE Transactions on Knowledge and Data Engineering, 30(9):1616–1637, 2018.

29. Emily Mastej, Lucas Gillenwater, Yonghua Zhuang, Katherine A Pratte, Russell P Bowler, and Katerina Kechris. Identifying protein–metabolite networks associated with copd phenotypes. Metabolites, 10(4):124, 2020.

30. Mikhail Belkin and Partha Niyogi. Laplacian eigenmaps for dimensionality reduction and data representation. Neural computation, 15(6):1373–1396, 2003.

31. Paul Erdos, Alfréd Rényi, et al. On the evolution of random graphs. Publ. Math. Inst. Hung. Acad. Sci, 5(1):17–60, 1960.

32. Gabor Csardi, Tamas Nepusz, et al. The igraph software package for complex network research. InterJournal, complex systems, 1695(5):1–9, 2006.

33. Patrick Danaher, Pei Wang, and Daniela M Witten. The joint graphical lasso for inverse covariance estimation across multiple classes. Journal of the Royal Statistical Society: Series B (Statistical Methodology), 76(2):373–397, 2014.

34. John L Hankinson, John R Odencrantz, and Kathleen B Fedan. Spirometric reference values from a sample of the general us population. American journal of respiratory and critical care medicine, 159(1):179–187, 1999.

35. Michael H Cho, Merry-Lynn N McDonald, Xiaobo Zhou, Manuel Mattheisen, Peter J Castaldi, Craig P Hersh, Dawn L DeMeo, Jody S Sylvia, John Ziniti, Nan M Laird, et al. Risk loci for chronic obstructive pulmonary disease: a genome-wide association study and meta-analysis. The lancet Respiratory medicine, 2(3):214–225, 2014.

36. Larry Gold, Deborah Ayers, Jennifer Bertino, Christopher Bock, Ashley Bock, Edward Brody, Jeff Carter, Virginia Cunningham, Andrew Dalby, Bruce Eaton, et al. Aptamer-based multiplexed proteomic technology for biomarker discovery. Nature Precedings, pages 1–1, 2010.

37. W Jenny Shi, Yonghua Zhuang, Pamela H Russell, Brian D Hobbs, Margaret M Parker, Peter J Castaldi, Pratyaydipta Rudra, Brian Vestal, Craig P Hersh, Laura M Saba, et al. Unsupervised discovery of phenotype-specific multi-omics networks. Bioinformatics, 35(21):4336–4343, 2019.

38. Wei Sun, Katerina Kechris, Sean Jacobson, M Bradley Drummond, Gregory A Hawkins, Jenny Yang, Ting-huei Chen, Pedro Miguel Quibrera, Wayne Anderson, R Graham Barr, et al. Common genetic polymorphisms influence blood biomarker measurements in copd. PLoS genetics, 12(8):e1006011, 2016.

39. Mark DM Leiserson, Fabio Vandin, Hsin-Ta Wu, Jason R Dobson, Jonathan V Eldridge, Jacob L Thomas, Alexandra Papoutsaki, Younhun Kim, Beifang Niu, Michael McLellan, et al. Pan-cancer network analysis identifies combinations of rare somatic mutations across pathways and protein complexes. Nature genetics, 47(2):106–114, 2015.

40. Christos Dimitrakopoulos, Sravanth Kumar Hindupur, Luca Häfliger, Jonas Behr, Hesam Montazeri, Michael N Hall, and Niko Beerenwinkel. Networkbased integration of multi-omics data for prioritizing cancer genes. Bioinformatics, 34(14):2441–2448, 2018.

41. Taisong Jin, Jun Yu, Jane You, Kun Zeng, Cuihua Li, and Zhengtao Yu. Lowrank matrix factorization with multiple hypergraph regularizer. Pattern Recognition, 48(3):1011–1022, 2015.

42. Sourav S Bhowmick and Boon Siew Seah. Clustering and summarizing protein-protein interaction networks: A survey. IEEE Transactions on Knowledge and Data Engineering, 28(3):638–658, 2015.

43. Danai Koutra, U Kang, Jilles Vreeken, and Christos Faloutsos. Vog: Summarizing and understanding large graphs. In Proceedings of the 2014 SIAM international conference on data mining, pages 91–99. SIAM, 2014.

44. Kristen LeFevre and Evimaria Terzi. Grass: Graph structure summarization. In Proceedings of the 2010 SIAM International Conference on Data Mining, pages 454–465. SIAM, 2010.

45. Yuanyuan Tian, Richard A Hankins, and Jignesh M Patel. Efficient aggregation for graph summarization. In Proceedings of the 2008 ACM SIGMOD international conference on Management of data, pages 567–580, 2008.

46. Ning Zhang, Yuanyuan Tian, and Jignesh M Patel. Discovery-driven graph summarization. In 2010 IEEE 26th international conference on data engineering (ICDE 2010), pages 880–891. IEEE, 2010.

