## Supplementary Information for "NetSHy: Network Summarization via a Hybrid Approach Leveraging Topological Properties"

### S.1 Existing approaches

Fig. S1 illustrates the overview of existing network summarization approaches utilizing feature profile ($X_{n \times p}$) and/or network connectivity ($A_{p \times p}$).

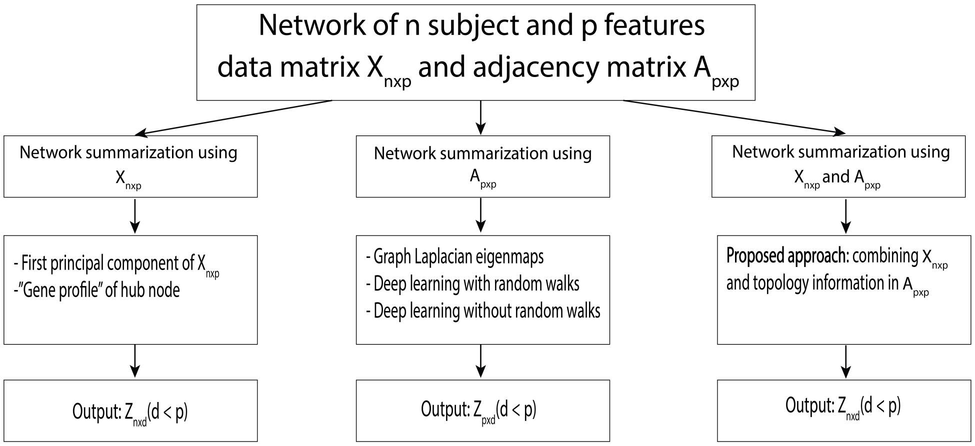


Fig. S1: **Overview of existing network summarization approaches:** Network summarization approaches utilizing only feature profiles $X_{n \times p}$ (left), only network connectivity $A_{p \times p}$ (middle), and both feature profiles $X_{n \times p}$ and network connectivity $A_{p \times p}$ (right); and their associated outputs $Z$.

### S.2 Additional information for the Materials and Methods section

#### COPD Phenotype

COPD was defined by spirometric evidence of airflow obstruction, which was computed as a ratio of post-bronchodilator forced expiratory volume at one second (FEV1) to forced vital capacity (FVC).

FEV1% is the amount of air one can forcibly exhale in one second (L) divided by the predicted FEV1 adjusted for age, height, race, and sex [8]. The Global Obstructive Lung Disease (GOLD) system is used to grade COPD. GOLD 0 represents an individual without COPD (FEV1 $\geq$ 80%; FEV1/FVC $>$ 0.7), GOLD 1 (FEV1 $\geq$ 80%; FEV1/FVC $<$ 0.7), GOLD 2 (50% $\leq$ FEV1 $<$ 80%; FEV1/FVC $<$ 0.7), GOLD 3 (30% $\leq$ FEV1 $<$ 50%; FEV1/FVC $<$ 0.7), and GOLD 4 (FEV1 $<$ 30%; FEV1/FVC $<$ 0.7), respectively represent the early, moderate, severe, and very severe stages of COPD.

**Proteomic data**

The following two platforms were used to quantify proteomic data in Visit 2 of COPDGene. *SOMAScan v1.3:* P100 plasma was profiled using SOMAscan® Human Plasma 1.3K assay (SomaLogic, Boulder, Colorado, CO, USA) at National Jewish Health. SOMAScan is a multiplex proteomic assay quantified by microarrays. This assay measured 1317 SOMAmers. SOMAmers are short single-stranded deoxyoligonucleotides (aptamers) that bind with high affinity and specificity to specific protein structures [7]. SomaLogic conducted quality assurance on each sample and normalized (hybridization and median), SOMAmers were calibrated (to remove inter-assay variation by analyte), and plate scaled (to adjust for total signal difference from plate to plate variation). As a final step, proteomic data was natural log transformed and standardized.

*SOMAScan v4.0:* The protein levels of EDTA (ethylenediaminetetraacetic acid) plasma samples from Visit 2 were quantified using SomaScan v4.0 (Somalogic, Boulder, Colorado). This SOMAScan platform used 4,979 different SOMAmers (aptamers) to quantify 4,776 unique proteins with 4,720 unique Uniprot numbers. SomaLogic conducted the following normalization and calibration to the results: signal normalization included plate hybridization to control for variability across array signals, median signal normalization to control for technical variability of replicates within a run, and plate scaling and calibration of SOMAmers to control for inter-assay variation between analytes and batch differences between plates. Then, median normalization to a reference using adaptive normalization by maximum likelihood was applied within the dilution group to quality control replicates and individual samples to remove edge effects and technical variance. Finally, the proteomic data was standardized.

**Metabolomic data**

The same P100 plasma was profiled using the Metabolon (Durham, NC, USA) Global Metabolomics platform. Briefly, untargeted gas chromatography–mass spectrometry and liquid chromatography–mass spectrometry (GC–MS and LC–MS) were used to quantify 1392 metabolites. A data normalization step was performed to correct variation resulting from instrument inter-day tuning differences: metabolite intensities were divided by the metabolite run day median, then multiplied by the overall metabolite median. It was determined that no further normalization was necessary based on the reduction in the significance of association between the top PCs and sample run day after normalization. Subjects with aggregate metabolite median z-scores greater than 3.5 standard deviation from the mean (n = 6) of the cohort were removed. Metabolites were excluded if more than 20% of samples were missing values [1]. For the 995 remaining metabolites, missing values were imputed across metabolites with k-nearest neighbors imputation (k = 10) using the R package "impute" [16]. As a final step, metabolomic data was natural log transformed and standardized.

**GWAS analysis**

We conducted the GWAS analysis on the University of Michigan Encore server (`https://github.com/statgen/encore`) using "Efficient and parallelizable association container toolbox" (EPACTS, `https://genome.sph.umich.edu/wiki/EPACTS`). Briefly, EPACTS efficiently performed statistical tests between phenotypes and sequence data through a user-friendly interface. We used TOPMed Freeze 10 (Jan 2022) whole genome sequence data from assembly GRCh38, a minor allele frequency (MAF) $> 0.1\%$, and a significance threshold of $5 \times 10^{-8}$ to perform these tests [18].

### S.3 Simulation Scenarios for Evaluation

Fig. S2 summarizes three simulation scenarios used to evaluate the proposed method NetSHy (in Section 2.2). Scenario (1) generated random graphs following Renyi-Erdos [5], provided a number of nodes $p$ and a graph sparsity $\alpha_0 \in [0, 1]$. The edge weights $\{w_{kl}\}_{k,l=1}^{p}$ connecting nodes $k$ and $l$ were simulated from the uniform distribution such that $w_{kl} \sim \text{Unif}(0.1, 0.8)$. Three network sizes of $p = 30, 60, \text{and } 100$, and three levels of sparsity $\alpha_0 = 0.3, 0.6, \text{and } 0.9$ were included in the study. Differently, Scenario (2) obtained the adjacency matrix $A$ directly from a previously published metabolite-protein network by Mastej et al. [12]. The network size and sparsity level were fixed at $p = 20$ and $\alpha_0 = 0.51$, respectively. However, we were able to additionally assess the impact of network sparsity at $\alpha_0 = 0.25$ by applying hard-thresholding to remove weak edges. Scenario (3) utilized the biological networks specific for chronic obstructive pulmonary disease (COPD) to validate the applicability of NetSHy. In this scenario, the observed data $X$ and phenotype $Y$ were used directly without any simulation involved.

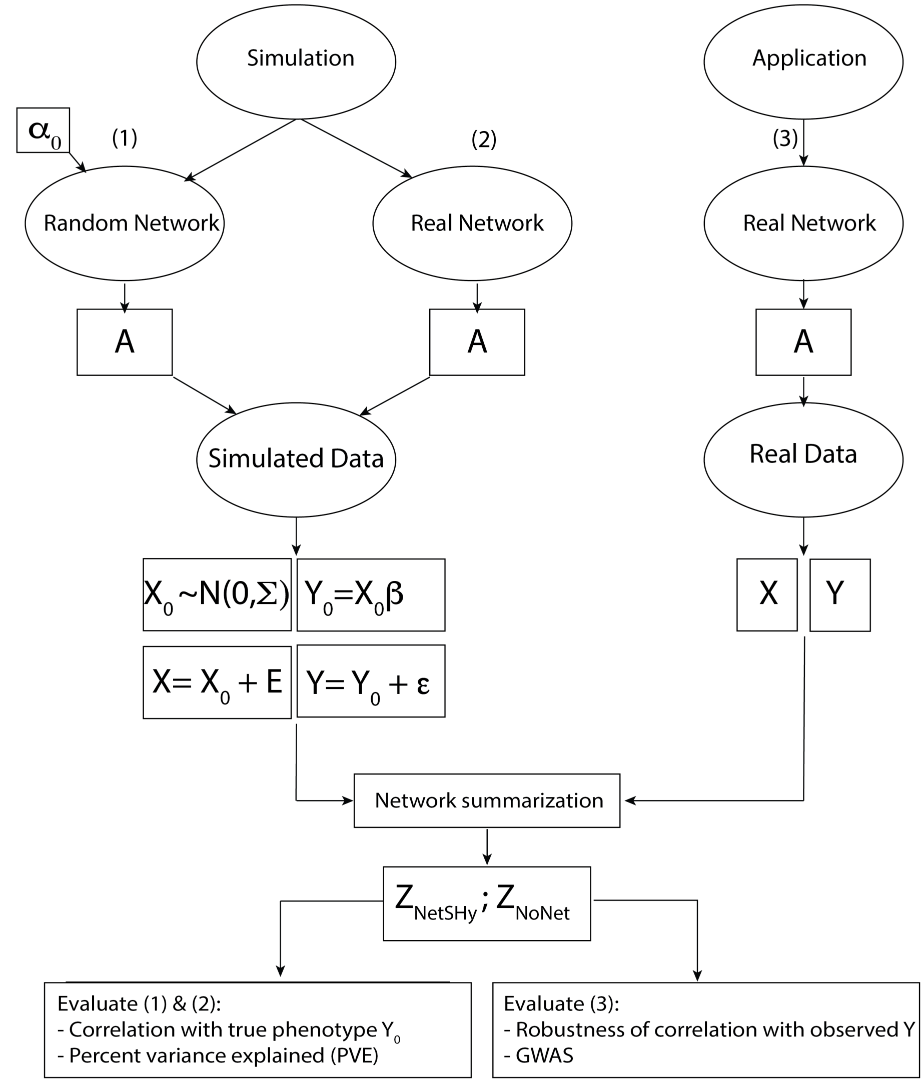


Fig. S2: **Scenario Setups for Evaluation.** Simulation Scenario (1): A network of size $p$ and sparsity level $\alpha_0$ generated from the Renyi-Erdos model, represented by an adjacency matrix $A$. Feature data matrix $X_0$ simulated from $A$ under Gaussian graphical model assumption, and Phenotype vector $Y_0$ generated as a linear function of $X_0$. Simulation Scenario (2): Directly utilized a previously published metabolite-protein network by Mastej et al. [12], represented by an adjacency matrix $A$. Steps to generate feature data matrix $X_0$ and phenotype vector $Y_0$ were same as in Scenario (1). Performances of the two approaches were evaluated based on correlation with true phenotype and PVE as in Section 2.2. Application (3): Directly used published metabolite-protein network [12] represented by an adjacency matrix $A$, and corresponding observed data matrix and phenotype vector as $X$ and $Y$, respectively. In this case, without knowledge of true underlying data matrix $X_0$, proportion of variance explained (PVE) was not assessed. Instead, the robustness of each approach regarding observed correlation with phenotype was of interest as the sample size decreased.

### S.4 Additional results of Simulation Scenario (1)

Figures S3 and S4 illustrate the performances of NetSHy and NoNet summarization scores at network sizes $p = 60$ and $p = 100$, respectively, in conjunction with increasing sparsity level $\alpha_0$, under Simuluation Scenario (1) in Section 2.2. Specifically, in each figure, the evaluation criteria were correlation ($\rho$) with true phenotype (top row) and proportion of variance explained (PVE) in true feature profile $X_0$ (bottom row) with respect to the optimal quantities $\rho_{opt}$ and $\text{PVE}_{opt}$, respectively. The closer the values to 1, the better the performances. Similar trends were observed across the two network sizes that there was no apparent difference between NetSHy and NoNet with regard to the true correlation $\rho$ with phenotype as sample size decreased. On the other hand, NetSHy yielded higher PVE in comparison with NoNet regardless of network sparsity, though the deviation in PVEs between the two approaches became less apparent at a larger $\alpha_0$. For instance, $\alpha_0 = 0.3$ and $n = 50$, the ratio of $\text{PVE}_{NetSHy}$ to the optimal $\text{PVE}_{opt}$ was 0.80 while such ratio of NoNet was 0.66. When the sparsity increased to $\alpha_0 = 0.9$, the PVEs of NetSHy and NoNet with respect to the optimal PVE were 0.67 and 0.62, respectively.

### S.5 GWAS result interpretation

The twenty-four significant SNPs obtained using NetSHy summary score fall in a region of the genome that is downstream from the gene pregnancy zone protein (PZP, Chromosome 12: 9,140,730-9,208,395), upstream of the alpha-2-macroglobulin pseudogene 1 (A2MP1) and overlaps with the gene killer cell lectin like receptor G1 (KLRG1; Chromosome 12: 8,950,044-9,215,657; `https://www.genecards.org`). PZP inhibits the activity of four classes of proteinases (serine, cysteine, aspartyl, and metalloproteases) and is highly expressed in late-pregnancy serum. It has been associated with phenotypes such as lymphocyte count and adolescent idiopathic scoliosis. While PZP is typically known to help prevent fetal rejection during pregnancy, it has also been associated with bronchiectasis, a chronic respiratory disease, in a study set in a specialist bronchiectasis clinic at Ninewells Hospital, Dundee, United Kingdom [6]. In the same vein, Smith et al. found an association of increased PZP in patients' sputum with the bronchiectasis severity index and FEV1% predicted [15]. Similarly, Mahor et al. named PZP as one of the proteins with altered expression in COPD patients [11]. Furthermore, KLRG1 is highly expressed in lymph nodes and the spleen. Associated phenotypes for KLRG1 are cholesterol measurements and eosinophil counts. Studies of copy number variations such as deletions or duplications have implicated this region

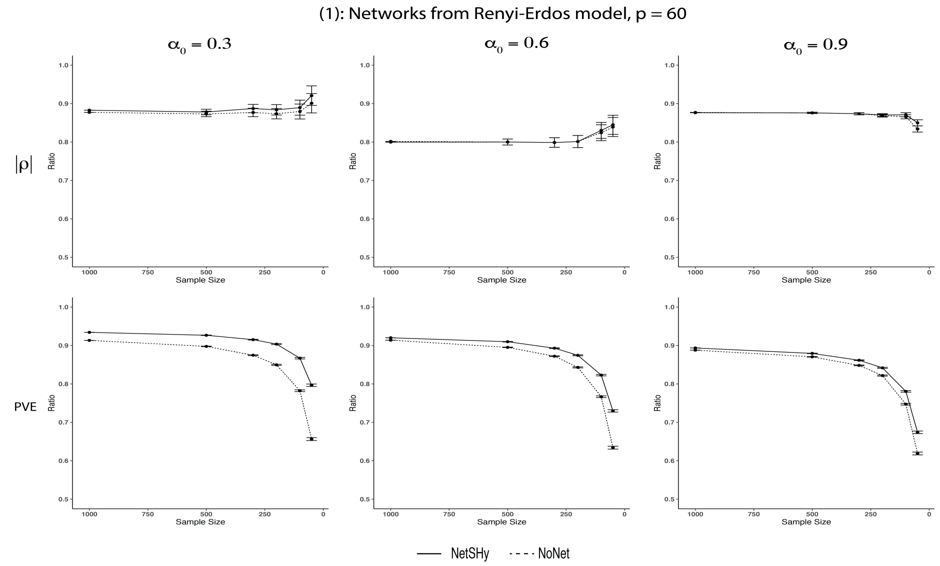


Fig. S3: **Results of Simulation Scenario (1), $p = 60$:** Fixing network size at $p = 60$ while varying sparsity levels from $\alpha_0 = 0.3$ to $\alpha_0 = 0.9$, NetSHy and NoNet were assessed using correlation with phenotype ($\rho$) and proportion of variance explained (PVE) relative to the optimal level, as sample size decreased. Specifically, the sample size was started at 1000 subjects, and random subsamplings were iterated 1000 times for each sample size of 500, 300, 200, 100, and 50, respectively. The closer the value to 1, the better the performance. The error bars summarize the standard deviations of $\rho$ and PVE from the 1000 iterations at each sample size excep for $n = 1000$. The range of $\rho$ and PVE ratio in the y-axis is between 0 and 1. However, we have zoomed in between 0.5 and 1 for better visualization.

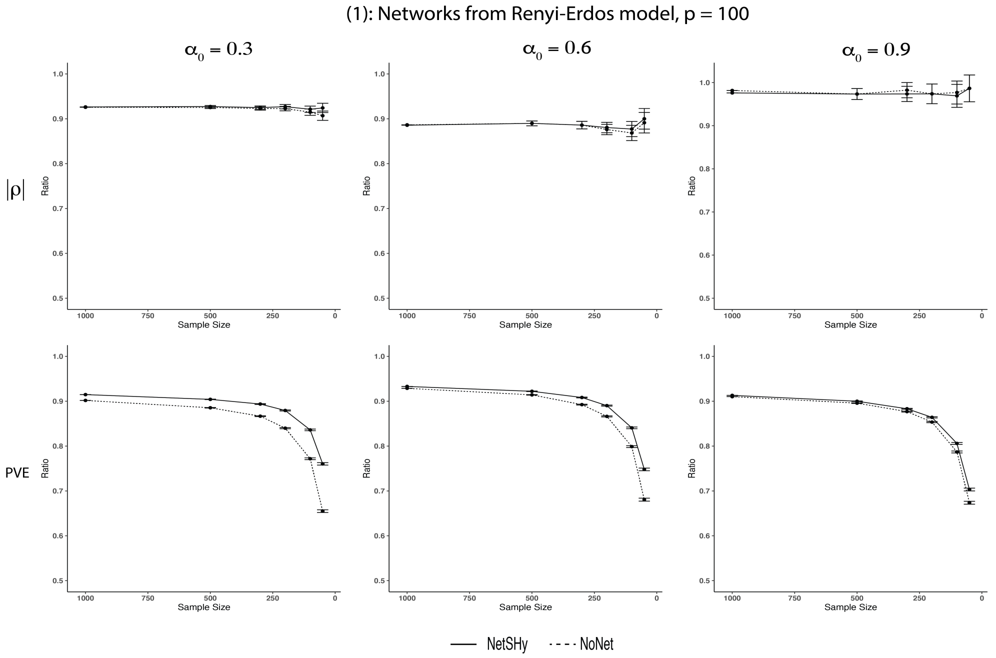


Fig. S4: **Results of Simulation Scenario (1), $p = 100$:** Fixing network size at $p = 100$ while varying sparsity levels from $\alpha_0 = 0.3$ to $\alpha_0 = 0.9$, NetSHy and NoNet were assessed using correlation with phenotype ($\rho$) and proportion of variance explained (PVE) relative to the optimal level, as sample size decreased. Specifically, the sample size was started at 1000 subjects, and random subsamplings were iterated 1000 times for each sample size of 500, 300, 200, 100, and 50, respectively. The closer the value to 1, the better the performance. The error bars summarize the standard deviations of $\rho$ and PVE from the 1000 iterations at each sample size excep for $n = 1000$. The range of $\rho$ and PVE ratio in the y-axis is between 0 and 1. However, we have zoomed in between 0.5 and 1 for better visualization.

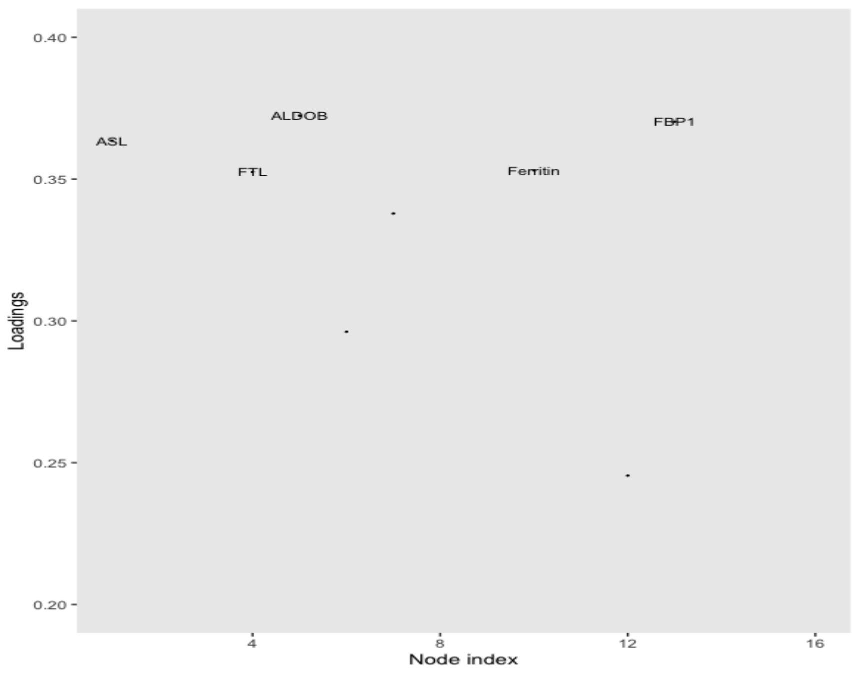


Fig. S5: **NetSHy weights**. Weights of five proteins which contribute the most to the NetSHy summary score. The five proteins include Fructose-bisphosphate aldolase B (ALDOB), Fructose-1,6-bisphosphatase 1 (FBP1), Argininosuccinate lyase (ASL), Ferritin, and Ferritin light chain (FTL).

(Band 12p13.31) as potentially pathogenic [3, 10, 2]. The top SNP rs118028480 (Chromosome 22: 39592172; [14] identified using NoNet score overlaps with the gene calcium voltage-gated channel subunit alpha1 I (CACNA1I; Chromosome 22: 39,570,753-39,689,737). The encoded protein for this gene is a "member of a low-voltage activated T-type calcium channel family and may be involved in calcium signaling in neurons" [14]. CACNA1I has been associated with intellectual disabilities and childhood epilepsy [14] and has been implicated in loss of function copy number variations [17].

Fig. S5 shows weights of five proteins which contribute the most to the NetSHy summary score including Fructose-bisphosphate aldolase B (ALDOB), Fructose-1,6-bisphosphatase 1 (FBP1), Argininosuccinate lyase (ASL), Ferritin, and Ferritin light chain (FTL). In a study performed by Pastor et al. [13], FBP1 was one of 40 identified proteins which were differentially expressed in lung cancer and COPD subjects as compared with the control group. Additionally, the level of iron-related protein like ferritin was shown to be higher in lung tissues of COPD individuals when compared to healthy controls, as shown by Zhang et al. [20].

### S.6 Additional analyses

We explored a diffusion process introduced by Leiserson et al. [9] and Dimitrakopoulos et al. [4] to identify subnetworks in Pan-Cancer studies. Though this did not directly obtain a network quantitative measure for each subject as we desire, it was worth inspecting the heat diffusion matrix as an alternative approach to capture the topology of the interaction network surrounding a node. Specifically, the diffusion matrix $F$ was defined as:

$$F = \beta(I - (1-\beta)W)^{-1},$$

where the $(ij)$th element of $W$ was denoted as:

$$W_{ij} = \begin{cases} \frac{1}{deg(j)}, \text{if node i interacts with node j} \\ 0, \text{otherwise} \end{cases}$$

In other words, $W$ served as a normalized adjacency matrix of the network. Note that the degree of diffusion in the network, i.e., $\beta$, was not known in advance. Similar to [4], we assessed 100 values of $\beta$ ranging from 0.001 to 1 to evaluate the performance of this heat diffusion process. In particular, for a given $\beta$ value, we obtained the corresponding heat matrix $F_\beta$. We then pursued the same procedure as NetSHy such that the matrix $F_\beta$ replaced the Laplacian matrix $L$ in transforming the molecular profile matrix $X_{n\times p}$. Explicitly, we defined $\tilde{X}_\beta = XF_\beta$. The corresponding summarization score was the first principal component of the PCA on $\tilde{X}_\beta$, represented by $Z^\beta_{DP}$ of dimension $n \times 1$, which was then used to compute a correlation ($\rho^\beta_{DP}$) with the phenotype $Y$. That is, $\rho^\beta_{DP} = corr(Z^\beta_{DP}, Y)$. Note that when $\beta = 1$, $F_\beta$ became the identity matrix, i.e., $F_{\beta=1} = I$. Consequently, for $\beta = 1$ we had $\tilde{X} = X$ and $\rho^\beta_{DP} = \rho_{NoNet}$.

We applied this diffusion process on the metabolite-protein network in Section 2.3.5. Fig. S6 shows the distribution of the absolute values of $\rho^\beta_{DP}$ calculated using the aforementioned diffusion process across 100 values of $\beta$. We noticed that 83% of the correlation values were less than 0.2. The maximum correlation ($|\rho_{DP}| = 0.34$) was reached when $\beta = 1$. In other words, we did not observe a significant gain in using this diffusion process in network summarization as compared to an approach of not using any network information, i.e., NoNet.

Furthermore, in our preliminary analysis, we attempted a weighted approach which took into account a secondary proximity, i.e., a topology overlap matrix (TOM) [19] in addition to the first-order proximity reflected in the Laplacian matrix ($L$). TOM captured the commonality in the neighborhoods of any two nodes in the network. In other words, we transformed the feature space

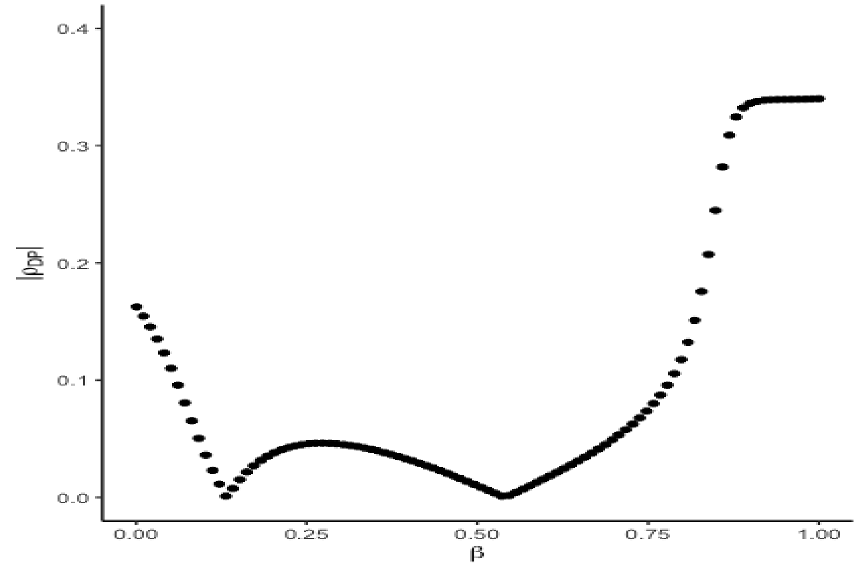


Fig. S6: **Correlation distribution using diffusion process**. Distribution of the absolute values of $\rho_{DP}^{\beta}$ calculated using the diffusion process across 100 values of $\beta$.

such that $X^* = (1-\alpha)X \times L + \alpha X \times TOM$ with $\alpha$ as a network density parameter. However, this weighted approach did not yield consistent and robust results in both simulation studies and real data analysis. As a result, we only focused on the first-order proximity captured in $L$ in this paper. With the lack of available methods that directly target our specific problem, the main focus of this manuscript was to introduce an approach which is effective in summarizing the network at a subject level while incorporating the topological properties. Altogether, we only compared NetSHy with the conventional approach utilizing only the node profiles, i.e. NoNet. As we briefly discussed in the Discussion Section, our future work would build upon NetSHy to explore different approaches to incorporate node profiles ($X$) and the network topology instead of utilizing the direct matrix multiplication.

### References


[1] Sabina Bijlsma, Ivana Bobeldijk, Elwin R Verheij, Raymond Ramaker, Sunil Kochhar, Ian A Macdonald, Ben Van Ommen, and Age K Smilde. Large-scale human metabolomics studies: a strategy for data (pre-) processing and validation. *Analytical chemistry*, 78(2):567–574, 2006.

[2] Bradley P Coe, Kali Witherspoon, Jill A Rosenfeld, Bregje WM Van Bon, Anneke T Vulto-van Silfhout, Paolo Bosco, Kathryn L Friend, Carl Baker, Serafino Buono, Lisenka ELM Vissers, et al. Refining analyses of copy number variation identifies specific genes associated with developmental delay. *Nature genetics*, 46(10):1063–1071, 2014.

[3] Gregory M Cooper, Bradley P Coe, Santhosh Girirajan, Jill A Rosenfeld, Tiffany H Vu, Carl Baker, Charles Williams, Heather Stalker, Rizwan Hamid, Vickie Hannig, et al. A copy number variation morbidity map of developmental delay. *Nature genetics*, 43(9):838–846, 2011.

[4] Christos Dimitrakopoulos, Sravanth Kumar Hindupur, Luca Häfliger, Jonas Behr, Hesam Montazeri, Michael N Hall, and Niko Beerenwinkel. Network-based integration of multi-omics data for prioritizing cancer genes. *Bioinformatics*, 34(14):2441–2448, 2018.

[5] Paul Erdos, Alfréd Rényi, et al. On the evolution of random graphs. *Publ. Math. Inst. Hung. Acad. Sci*, 5(1):17–60, 1960.

[6] Simon Finch, Amelia Shoemark, Alison J Dicker, Holly R Keir, Alexandria Smith, Samantha Ong, Brandon Tan, Jean-Yu Choi, Thomas C Fardon, Diane Cassidy, et al. Pregnancy zone protein is associated with airway infection, neutrophil extracellular trap formation, and disease severity in bronchiectasis. *American journal of respiratory and critical care medicine*, 200(8):992–1001, 2019.

[7] Larry Gold, Deborah Ayers, Jennifer Bertino, Christopher Bock, Ashley Bock, Edward Brody, Jeff Carter, Virginia Cunningham, Andrew Dalby, Bruce Eaton, et al. Aptamer-based multiplexed proteomic technology for biomarker discovery. *Nature Precedings*, pp. 1–1, 2010.

[8] John L Hankinson, John R Odencrantz, and Kathleen B Fedan. Spirometric reference values from a sample of the general us population. *American journal of respiratory and critical care medicine*, 159(1):179–187, 1999.

[9] Mark DM Leiserson, Fabio Vandin, Hsin-Ta Wu, Jason R Dobson, Jonathan V Eldridge, Jacob L Thomas, Alexandra Papoutsaki, Younhun Kim, Beifang Niu, Michael McLellan, et al.

Pan-cancer network analysis identifies combinations of rare somatic mutations across pathways and protein complexes. *Nature genetics*, 47(2):106–114, 2015.

[10] Irene Madrigal, Margarita Martinez, Laia Rodriguez-Revenga, Ana Carrió, and Montserrat Milà. 12p13 rearrangements: 6 mb deletion responsible for id/mca and reciprocal duplication without clinical responsibility. *American Journal of Medical Genetics Part A*, 158(5):1071–1076, 2012.

[11] Durga Mahor, Vandana Kumari, Kapil Vashisht, Ruma Galgalekar, Ravindra M Samarth, Pradyumna K Mishra, Nalok Banerjee, Rajnikant Dixit, Rohit Saluja, Sajal De, et al. Elevated serum matrix metalloprotease (mmp-2) as a candidate biomarker for stable copd. *BMC Pulmonary Medicine*, 20(1):1–9, 2020.

[12] Emily Mastej, Lucas Gillenwater, Yonghua Zhuang, Katherine A Pratte, Russell P Bowler, and Katerina Kechris. Identifying protein–metabolite networks associated with copd phenotypes. *Metabolites*, 10(4):124, 2020.

[13] María Dolores Pastor, Ana Nogal, Sonia Molina-Pinelo, R Melendez, Ana Salinas, M González De la Peña, J Martín-Juan, J Corral, Rocío García-Carbonero, Amancio Carnero, et al. Identification of proteomic signatures associated with lung cancer and copd. *Journal of proteomics*, 89:227–237, 2013.

[14] Stephen T Sherry, M-H Ward, M Kholodov, J Baker, Lon Phan, Elizabeth M Smigielski, and Karl Sirotkin. dbsnp: the ncbi database of genetic variation. *Nucleic acids research*, 29(1):308–311, 2001.

[15] Alexandria Smith, Jean-Yu Choi, Simon Finch, Samantha Ong, Holly Keir, Alison Dicker, and James Chalmers. Sputum pregnancy zone protein (pzp)-a potential biomarker of bronchiectasis severity, 2017.

[16] RT Trevor Hastie, R Tibshirani, B Narasimhan, and G Chu. impute: impute: Imputation for microarray data. *R Package*, 2018.

[17] Kendy K Wong, Ronald J deLeeuw, Nirpjit S Dosanjh, Lindsey R Kimm, Ze Cheng, Douglas E Horsman, Calum MacAulay, Raymond T Ng, Carolyn J Brown, Evan E Eichler, et al. A comprehensive analysis of common copy-number variations in the human genome. *The American Journal of Human Genetics*, 80(1):91–104, 2007.

[18] ChangJiang Xu, Ioanna Tachmazidou, Klaudia Walter, Antonio Ciampi, Eleftheria Zeggini, Celia MT Greenwood, and UK10K Consortium. Estimating genome-wide significance for whole-genome sequencing studies. *Genetic epidemiology*, 38(4):281–290, 2014.

[19] Bin Zhang and Steve Horvath. A general framework for weighted gene co-expression network analysis. *Statistical applications in genetics and molecular biology*, 4(1), 2005.

[20] William Z Zhang, Clara Oromendia, Sarah Ann Kikkers, James J Butler, Sarah O'Beirne, Kihwan Kim, Wanda K O'Neal, Christine M Freeman, Stephanie A Christenson, Stephen P Peters, et al. Increased airway iron parameters and risk for exacerbation in copd: an analysis from spiromics. *Scientific reports*, 10(1):1–13, 2020.
